# Electronic health data exploring cardiorespiratory responses of transfusions in preterm infants: An international multicenter cohort study

**DOI:** 10.64898/2026.09.01.26361418

**Authors:** Antoine Honoré, Till Rech, Alexandra Scrivens, Ivan Binotto, Coen S. Zandvoort, Hilde van der Staaij, Mariska Peck, Sanja Zivanovic, Simon J. Stanworth, Caroline Hartley, Christof Dame, Emöke Deschmann, the Neonatal Transfusion Network

**Author notes:** **Corresponding Author:** Christof Dame, MD **Corresponding author email address:**. Contributed equally as co-first authors (Antoine Honoré, Till Rech). Contributed equally as co-senior authors (Caroline Hartley, Christof Dame, Emöke Deschmann). A complete list of study group members appears in the Acknowledgements.

## Abstract

**Background and Objectives:** Preterm infants are commonly transfused, yet direct cardiorespiratory effects of red blood cell (RBC) transfusions remain poorly understood. We explored the feasibility of using multicentre electronic health data (EHD) to study such cardiorespiratory responses.

**Methods:** Highly granular routine EHD were collected from preterm infants born <32 weeks gestational age at three European centres. Heart rate, oxygen saturation, and respiratory rate were evaluated 12 hours before and after the RBC transfusion.

**Results:** A total of 321 transfusions in 164 infants were analysed. Overall, there was no significant change in the rate of bradycardia and apnoea following transfusion. Cardiorespiratory parameters varied substantially between infants; e.g. 20% of transfusions were associated with an unexpected, significant increase in heart rate. Respiratory rate and oxygen saturation exhibited similarly heterogenous patterns following transfusion. In sub-group analysis, the proportion of transfusions with increased heart rate was significantly higher within the first two weeks than later (32% vs 13%, p=0.0019).

**Conclusions:** Multicentre EHD extraction allows to identify otherwise masked short-term effects of RBC transfusions on cardiorespiratory parameters, possibly indicating cardiac or pulmonary overload. Such effects may vary with adaptation to anaemia. Analysing EHD may ultimately enable personalized transfusion practice.

## INTRODUCTION

Red blood cell (RBC) transfusion is a common intervention in the neonatal intensive care of very preterm infants, who frequently develop anaemia.^1, 2^ Although perceived as a standard, often life-saving and low-risk intervention, RBC transfusions may be associated with increased incidence of morbidities such as necrotizing enterocolitis (NEC), bronchopulmonary dysplasia (BPD) and retinopathy of prematurity (ROP).^3–6^ Moreover, there may be under-recognition of acute transfusion-related side effects. In adults, acute risks include transfusion-associated circulatory overload (TACO), transfusion-related acute lung injury (TRALI) and transfusion-related acute gut injury (TRAGI); the frequency of these risks in infants is unknown. In neonates, (single-unit) volumes for transfusion may be as high as 15 or 20 ml/kg, which is very high compared to that in adults. Thus, RBC transfusions have the potential to disturb the haemodynamics and respiration of the neonate. It is important to characterize the complex physiological responses to transfusion in order to weight the risks versus benefits of the intervention.

Previous studies provided inconsistent results regarding neonatal cardiorespiratory responses to transfusion, with some studies demonstrating reduction in apnoea rate following transfusion and others not identifying such effects.^7–11^ Recently, a secondary analysis of the TOP (Transfusion of Prematures) randomized clinical trial found that RBC transfusion was associated with increased regional oxygenation of both brain and mesenteric tissue despite no change in peripheral oxygen saturation (SpO2).^12^ Another study showed that both the number of desaturations and the area under the curve of SpO2 concentrations less than 80% decreased after RBC transfusion.^7^ These studies have largely focused on group average effects, however, it is unclear if some individuals may have adverse cardiorespiratory responses to transfusion. Given that adverse effects may only occur in a small number of infants, analysis of a large sample size is essential to identifying such an effect.

The aim of this study was to explore the feasibility of using continuously monitored cardiorespiratory parameters in preterm infants born <32 weeks gestational age, using data obtained from multiple European centres, and to elucidate the cardiorespiratory impact of RBC transfusion. We hypothesized that data from multiple centres enables identification of infrequent cardiovascular effects of RBC transfusions in very preterm infants.

## METHODS

### Recruitment of study sites

Within the Neonatal Transfusion Network (https://neonataltransfusionnetwork.com/) four European academic centres with tertiary level neonatal intensive care units (NICUs) in Berlin, London, Oxford and Stockholm participated in the study. Data collection in Berlin has been delayed due to extensive regulatory and implementation requirements for data extraction from the electronic health data (EHD) platform and so no data was obtainable to date (June 2026) from this study centre.

### Study design and governance

Participating sites were responsible for obtaining all local and national regulatory approvals to establish relevant local data repositories. Each centre was responsible for maintenance and security of its institutional data. The study conformed to the standards set by the Declaration of Helsinki and Good Clinical Practice. The study was approved, or granted a waiver of consent, by the responsible institutional review board or research ethics committee at each participating site (see Supplemental Information). The study is reported in accordance with the STROBE guidelines for observational studies.

Pathways for use of EHD were different at all participating sites. Detailed information on the access to electronic health records in the single centres is provided in the supplements (supplemental figures S1.1–S1.4). The online supplemental material also summarizes electronic infrastructures and relevant linkages of all centres (supplemental table on monitoring systems). Due to local equipment, data collection was retrospective in London and Stockholm and prospective in Oxford.

### Inclusion criteria

Preterm infants born <32 weeks of gestation, who received one or more RBC transfusions during NICU admission and had continuous digital vital monitoring signs, were eligible for inclusion into the study database at each participating centre.

### Data extraction

#### Retrospective data extraction (London, Stockholm)

Continuous monitoring data were retrieved from each hospital’s data warehouse and linked to RBC- transfusion events. For each event we recorded start and end time, volume, weight-adjusted dose, rate and product specification; outlier events were removed (only necessary for Stockholm data, manual plausibility check in London, see Supplementary Methods). Each event was associated with the last pre-transfusion haemoglobin concentration (within 24 hours) and, where available, the last recorded mode of respiratory support before the event. Heart rate (HR) was derived from electrocardiogram-based interbit interval (IBI) signal, peripheral blood oxygen saturation (SpO2) from non-invasive pulse oximetry, and respiratory rate (RR) from chest impedance; per-centre sampling rates and monitoring/storage systems are summarized in the online supplemental material.

#### Prospective data collection (Oxford)

In Oxford, no vital-signs data warehouse existed, thus data was collected prospectively. Vital signs (HR, RR, SpO2; 1 Hz) were continuously downloaded from the bed-side monitor (interface computer solution) for at least 12 hours before and after the start of transfusion, and impedance pneumography (62.5 Hz) was additionally recorded, enabling accurate quantification of apnoeas. Transfusion details, pre-transfusion haemoglobin, respiratory support and demographics were obtained from electronic and paper records (see supplementary methods).

In all centres, SIPPV/SIMV and invasive HFO were classed as mechanical ventilation; the full range of respiratory-support modes encountered is listed in the Supplementary Methods.

### Data analysis

Analysis was performed in MATLAB (MathWorks) separately for each centre. Data were federated: raw data were analysed within each centre and only pre-processed output was combined; owing to ethical restrictions, Stockholm data did not leave the centre.

Analysis steps are illustrated exemplarily for single infants in supplemental figure S2. Data were epoched from 12 hours before to 12 hours after the start of transfusion and aligned to its onset. For HR, RR and SpO2, we computed both the mean and the standard deviation (variability) in 1-hour windows advanced in 30-minute steps. Transfusion events without a minimum of 3 hours data recording within the 6 hours before the start of transfusion and 6 hours of data recording within the 12 hours after initiation of the transfusion were removed from the analysis.

#### Identifying physiological response to RBC transfusion

Parameters were baseline-corrected by subtracting the pre-transfusion mean. For each transfusion event and parameter, a post-transfusion change was considered significant when the value remained beyond one standard deviation of the baseline for at least 6 consecutive time points (at least 3 hours). The basis for this threshold is given in the Supplementary Methods.

#### Identifying cardiorespiratory events

Cardiorespiratory events comprised oxygen desaturations, apnoeas and bradycardias, identified using established criteria^13, 14^; for thresholds and full definitions see the Supplementary Methods. Event rates were computed per hour, separately for each event type, averaged before and after transfusion.

### Statistical analysis

Rates of cardiorespiratory events before and after transfusion were compared using Wilcoxon signed-rank tests. Changes in cardiorespiratory parameters were assessed with cluster-based permutation tests;^15^ test details, including the permutation procedure and software,^16^ are given in the Supplementary Methods.

Differences between centres in the proportion of changes in each cardiorespiratory parameter were assessed using Fisher’s exact test. In an exploratory analysis, to compare early versus late transfusions and their effect on cardiorespiratory parameters, permutation tests were used due to the small number of transfusion events in some groups. As this was conducted as an exploratory analysis, p-values are presented only as a guide. Finally, two-proportion z-tests were used to compare percentages of transfusion events with an increase in mean HR and a decrease in mean RR in the infants who were younger than 2 weeks and in those greater than or equal to two weeks postnatal age.

### Code availability

Code for data analysis is available from GitHub (https://github.com/antoinehonore/neored-vs).

## RESULTS

### Data flows and study participants

We established the extraction of EHD at independent European tertiary level NICUs and successfully obtained high frequency cardiorespiratory data from routine monitoring linked to RBC transfusion. However, there were challenges: Data collection in Berlin was not completed in time for inclusion (see Methods). Similarities and differences between centres in access to the data are illustrated in the supplemental material (supplemental figures S1.1-S1.4 and table S1).

In total, the study included 164 infants who received 321 RBC transfusions during neonatal intensive care (figure 1). Among them, 298 transfusions were recorded from repositories that store routine monitoring data (data warehouses in London [88 transfusions in 44 infants] and Stockholm [210 transfusions in 97 infants]). Another 23 transfusions were prospectively recorded in 17 infants. Demographic and transfusion data are summarized in table 1.

**Figure 1.**
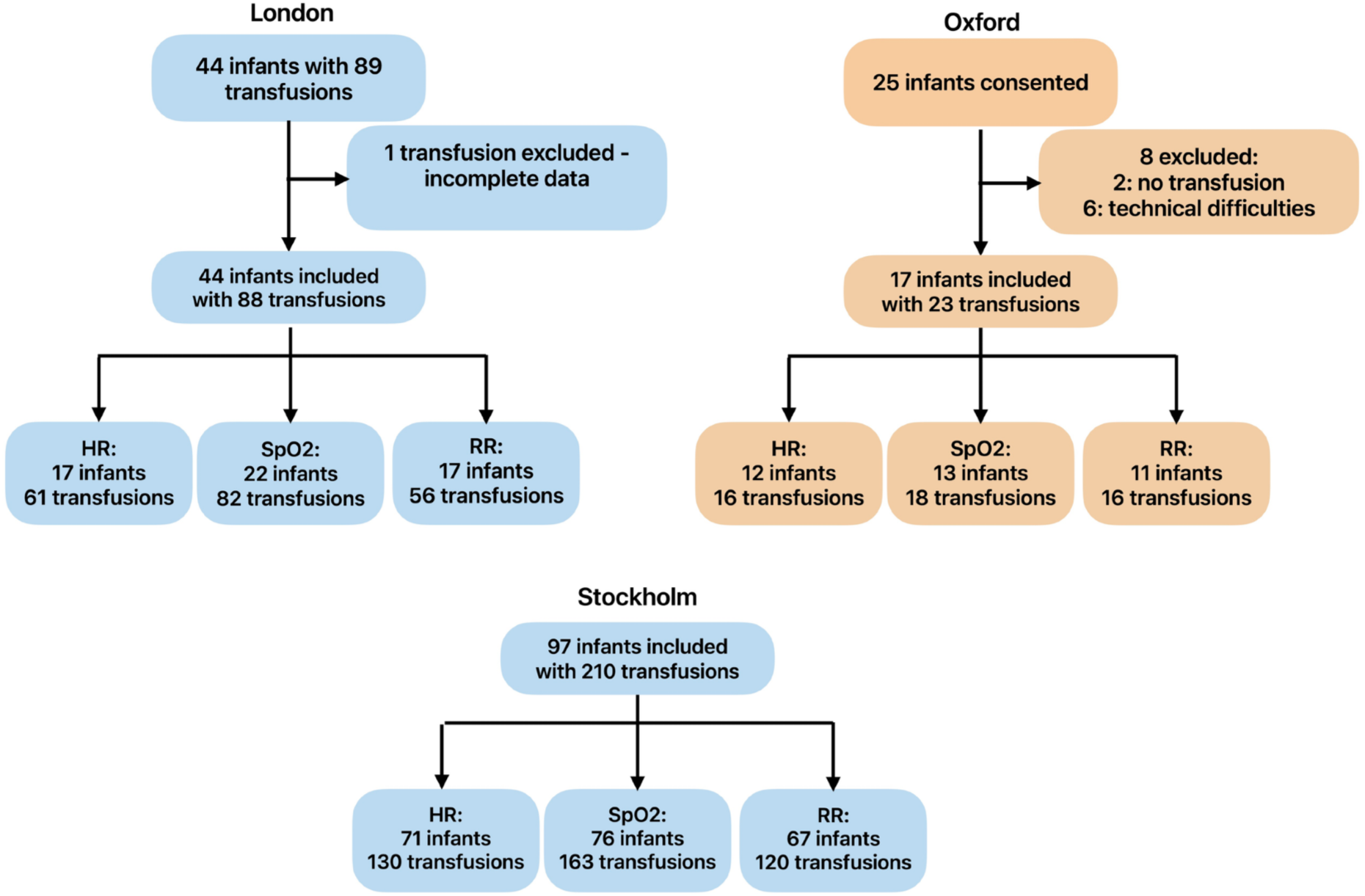
Participant flowchart. Number of patients available for the analysis of RBC transfusion effects on routine vital-sign monitoring parameters in each study centre. Blue boxes: retrospective data collection; orange boxes: prospective data collection.

**Table 1.** Demographic and transfusion characteristics of the study populations. Data are presented as n (%) or median (lower quartile, upper quartile).

|  | London | Oxford | Stockholm |
| --- | --- | --- | --- |
| **Demographics** |  |  |  |
| Patients (n) | 24 | 17 | 97 |
| Sex (female), n (%) | 13 (54) | 3 (18) | 49 (51) |
| Gestational age at birth (weeks) | 24.79 (24–25.39) | 25.14 (24.57–27.14) | 26.57 (25.29–27.29) |
| Birth weight (g) | 677.5 (573–863.25) | 760 (640–795) | 786 (636–966) |
| <750 g, n (%) | 13 (54) | 7 (41) | 41 (42) |
| ≥750 to 1000 g, n (%) | 9 (38) | 8 (47) | 33 (34) |
| ≥1000 to 1500 g, n (%) | 2 (8) | 2 (12) | 20 (21) |
| **Transfusion management** |  |  |  |
| RBC transfusions analysed (n) | 88 | 23 | 210 |
| Postconceptional age at transfusion (weeks) | 27.29 (25.29–29.29) | 29.43 (27.57–32.14) | 29.35 (27.64–31.39) |
| Postnatal age at transfusion (weeks) | 2.14 (0.71–4.71) | 4.00 (3.14–5.42) | 2.32 (0.65–4.60) |
| Pre-transfusion haemoglobin (g/L), within 24 h | 94.5 (87–104) | 86.0 (81.0–89.5) | 105.5 (96.0–114.0) |
| Transfusion volume (ml/kg) | 20 (20–20) | 14.55 (13.00–18.10) | 13.45 (12.51–13.82) |
| Transfusion rate (ml/kg/h) | 5 (5–5) | 3.59 (3.27–4.18) | 4.55 (4.26–5.00) |
| Mechanical ventilation at start, n (%) | 67 (76) | 9 (39) | 87 (41) |
| Non-invasive ventilatory support at start, n (%) | 21 (24) | 14 (61) | 123 (59) |

There was considerable variation between centres in the age of infants included, pre-transfusion haemoglobin concentration (differing by up to 20 g/L), transfusion volume and rate (>30% variation), and the proportion receiving invasive versus non-invasive respiratory support (table 1).

### Effects of RBC transfusion on the rate of oxygen desaturations, apnoea and bradycardia

Overall, there was a small, but significant, decrease in the rate of oxygen desaturations following transfusion (figure 2A, Stockholm: median difference = 0.08 desaturations per hour, P = .0016; Oxford: median difference = 0.25 desaturations per hour, P = .063, Wilcoxon signed rank test; note not assessed in the London data as the granularity [sampling rate] was lower than the minimum desaturation duration). There was no significant difference in the rate of bradycardia (figure 2B, Stockholm: P = .36, Oxford: P = .23, Wilcoxon signed rank test, London: not assessed). However, there was a trend towards a decrease in apnoea rate (figure 2C, Oxford: P = .091; in non-ventilated infants only: P = .35; note that apnoea was only measured in the Oxford data – see methods).

**Figure 2.**
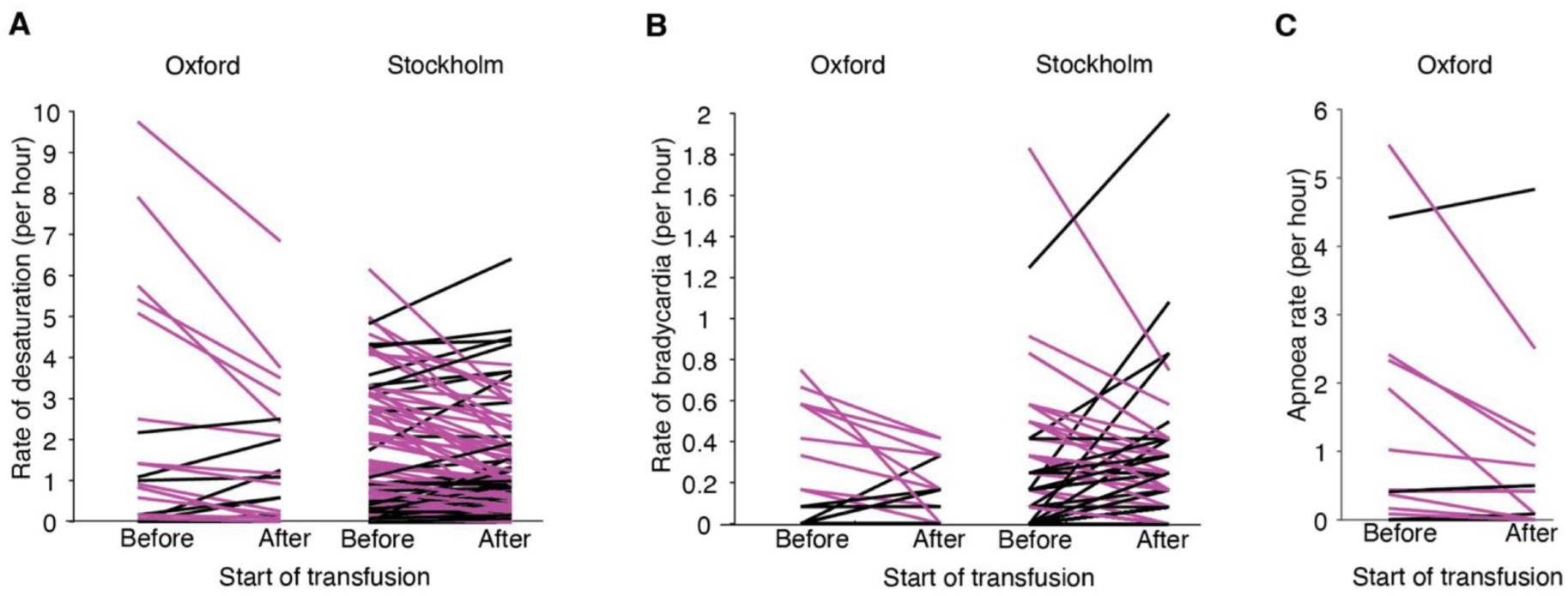
Group-level cardiorespiratory effects of RBC transfusion. Rate of oxygen desaturations (A; SpO2 <80%, ≥15s), bradycardias (B; <100bpm, ≥ 10s) and apnoeas (C; ≥15s) in the 12 hours before and after the start of transfusion. Purple = decrease, black = no change/ increase. Desaturations and bradycardias assessable in Oxford and Stockholm (owing to sampling rate); apnoeas only in the Oxford, where impedance pneumography was recorded, enabling accurate quantification (see Methods).

### Effects of RBC transfusion on cardiorespiratory parameters of preterm infants

Overall, averaging across all transfusions at each centre, there was relatively little change in cardiorespiratory response following transfusion, with a small but significant decrease in SpO2 variability following transfusion observed at all centres (P = .001, Oxford & Stockholm, P = .009, London, cluster-based permutation test), a significant decrease in mean heart rate in the Stockholm data only (P = .004), and no significant differences in other cardiorespiratory parameters at the other group levels.

In contrast, some individual infants displayed clear changes in cardiorespiratory monitoring data in response to transfusion (examples in figure 3A). Overall, 28% of transfusion events showed a significant decrease in mean HR after initiation of RBC transfusion (at least one SD below the pre-transfusion baseline for at least 3 hours), 19% a significant increase, and 53% no change (figure 3B). Similarly, mean RR remained stable in 58% of transfusions (decreased in 24%, increased in 18%).

**Figure 3.**
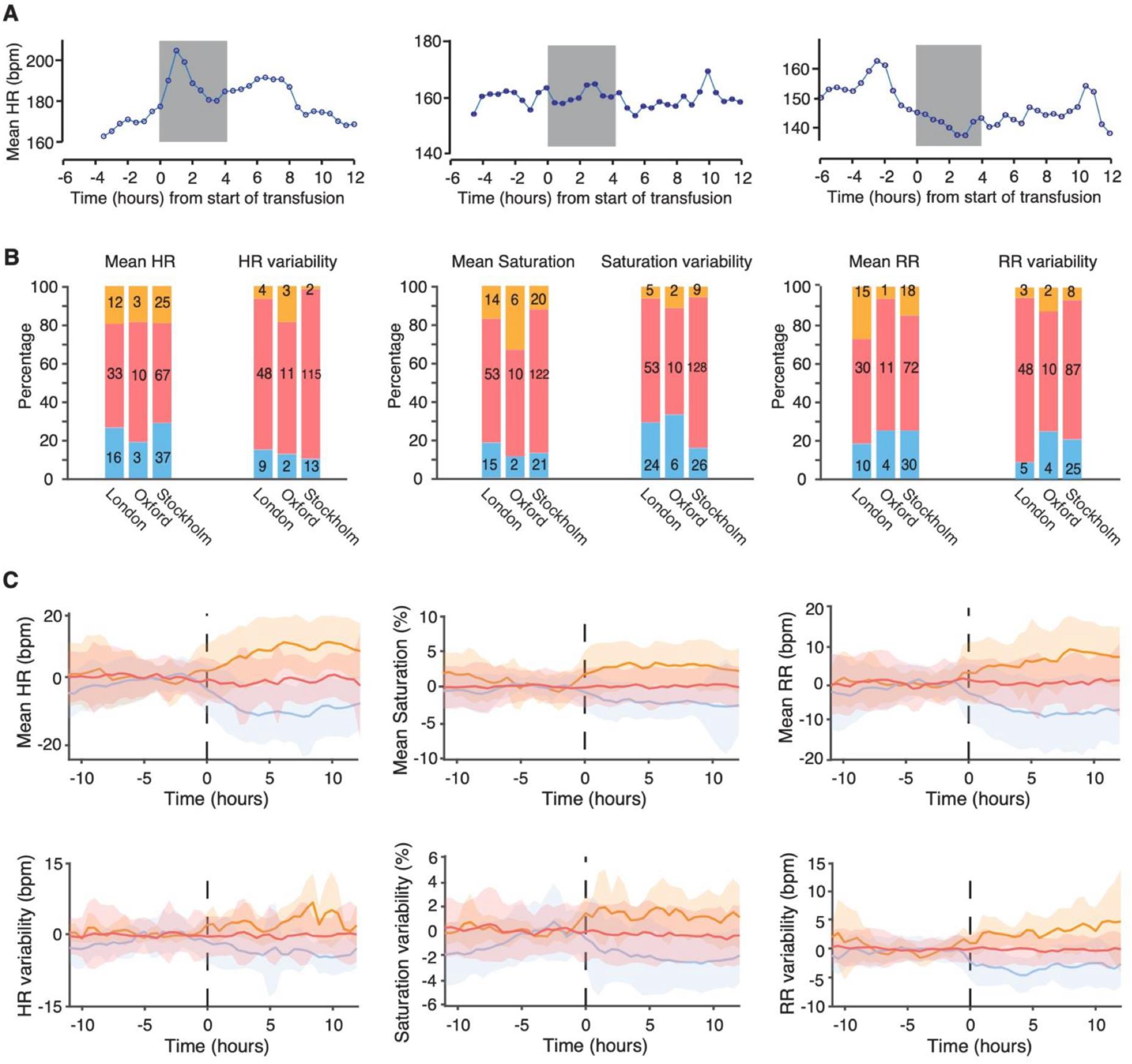
Variability in the cardiorespiratory response to transfusion. (A) Example infants showing an increase, no change, decrease in mean HR after start of transfusion (left to right). (B) Distribution of transfusion response groups in each cardiorespiratory metric, by centre. Numbers within bars indicate the absolute number of transfusion events. Red: no change; blue: decrease; orange: increase. (C) Longitudinal HR, RR and saturation (mean and variability) for increase- (orange), decrease- (blue) or no change- (red) subgroups combined across centres. HR, heart rate (bpm); RR, respiratory rate (breaths/min); saturation, peripheral oxygen saturation (%).

Mean SpO2 remained stable in 70% (decreased in 15%, increased in 15%; figure 3B).

There was no significant difference in the proportion of transfusion events exhibiting changes in the cardiorespiratory parameters between centres (P > .1) except for HR variability (P = .019, Fisher’s exact test), with comparatively low numbers of transfusions in Stockholm showing an increase in HR variability, and SpO2 variability (P = .040, figure 3B). The federated dataset confirmed the major patterns in each cardiorespiratory parameter in response to the RBC transfusion (figure 3C).

### Different cardiorespiratory response to early vs late RBC transfusion in preterm infants

In an exploratory analysis of factors affecting the cardiorespiratory response, postnatal age, pre-transfusion haemoglobin concentration and postmenstrual age were compared between transfusion events with no change, a decrease, or an increase in a parameter (figure 4A–F). For mean HR, there were significant differences by postnatal age (P = .018), pre-transfusion haemoglobin (P = .015) and postmenstrual age (P = .036); a difference in postnatal age was also seen relative to the mean RR response (P = .049). Group medians are shown in figure 4. Grouping transfusion events according to whether they occurred in the first two weeks of life (early anaemia) or from 2 weeks of life onwards (late anaemia), the proportion of infants who exhibited an increase in mean HR was significantly higher in the infants in the early anaemia group compared to those in the late anaemia group (32% compared with 13%, P = .0019, two-proportion z-test, figure 4G). The proportion of infants who exhibited a decrease in mean RR was significantly higher in the early anaemia group (33%) compared with 19% in the late anaemia group (P = .038, figure 4H).

**Figure 4.**
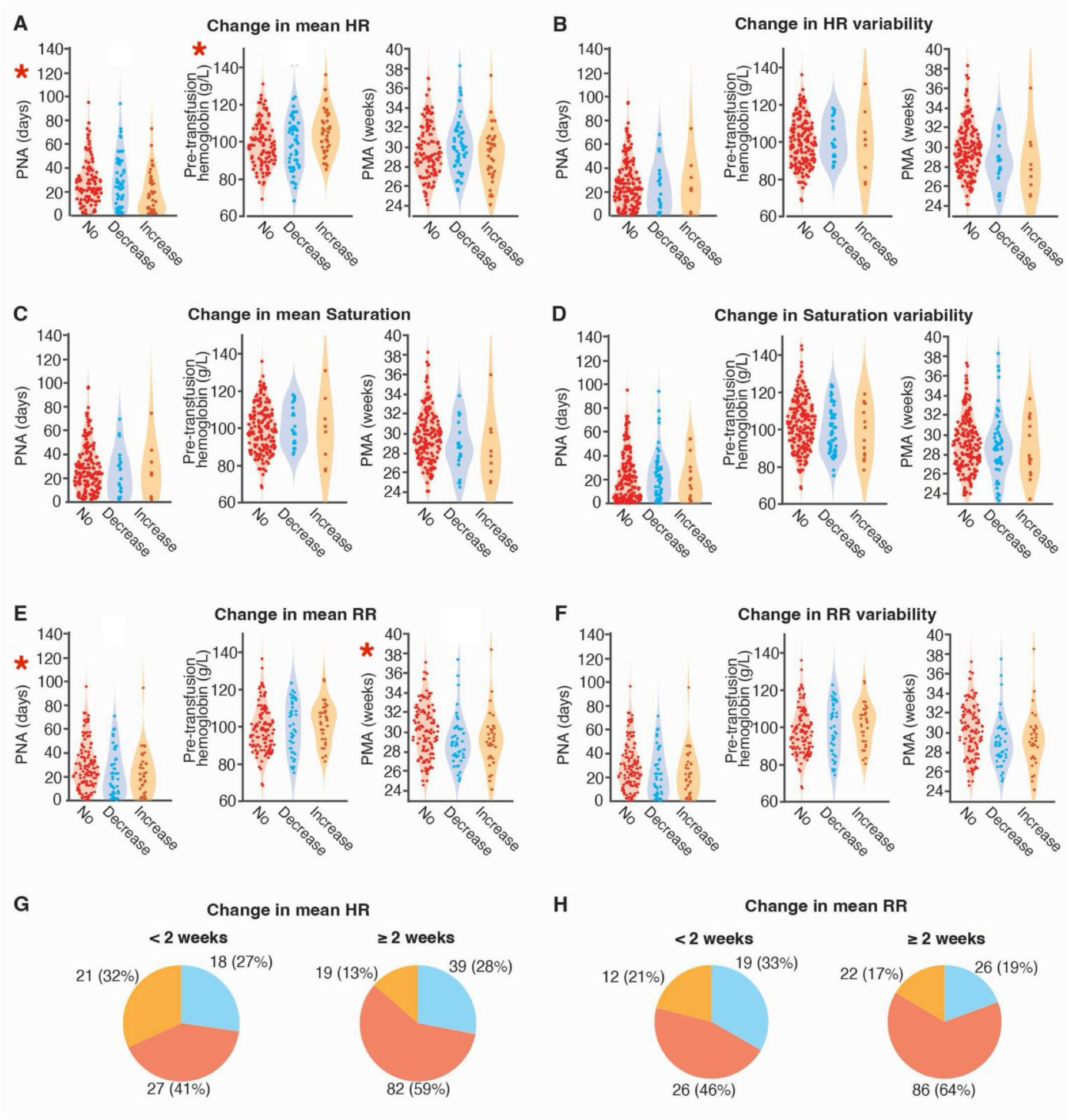
Variability in response may be related to age and pre-transfusion haemoglobin. (A–F) Violin plots of postnatal age (PNA), pre-transfusion haemoglobin and postmenstrual age (PMA) by response group. Red: no change; blue: decrease; orange: increase. Each dot = one transfusion event; centres combined. Asterisk: P <.05 (permutation test) for the comparison across groups. (G, H) Distribution of transfusion events with no change (red), decrease (blue) or increase (orange) in mean HR (G) and mean RR (H), by PNA (<2 vs. ≥2 weeks) at transfusion. Numbers = transfusion events, data combined across centres. HR, heart rate; RR, respiratory rate; saturation, peripheral oxygen saturation.

## DISCUSSION

We established the feasibility of extracting and federating EHD from NICUs across independent European centres to explore changes in cardiorespiratory parameters related to RBC transfusion. The analysis of individual cardiorespiratory responses demonstrated that some infants exhibit marked changes in cardiorespiratory parameters. The combined data indicate that these changes may be related to postnatal age and pre-transfusion haemoglobin, with infants receiving RBC transfusion(s) in the first two weeks of life more likely to have a significant increase in mean HR following transfusion. Thus, our study extends work by Poppe et al,^7^ by evaluating individual patient level data. Consistent with previous data,^7, 17, 18^ we observed a small but significant decrease in the rate of oxygen desaturations following transfusion, and a significant decrease in oxygen saturation variability, likely mediated through an enhanced oxygen carrying capacity. The reasons for, and clinical impact of the observed changes following transfusion remain unknown. The severity and duration of oxygen desaturations may increase the risk of disorders such as BPD, ROP and poor long-term neurodevelopmental outcomes,^19–21^ and argue for a concept of haemovigilance that considers the impact of transfusion on cardiorespiratory parameters. Adult donor blood components could induce storage lesions or release of biological response modifiers and so an increased HR might suggest an inflammatory response.^22, 23^ However, effects of RBC transfusion on cardiac output and volume overload should also be considered. Other research has indicated that preterm infants (<32 weeks of gestation) with pretransfusion haematocrit levels <27% had higher left ventricular end systolic and diastolic diameters in comparison to those with haematocrit levels ≥27%.^24^ After transfusion, peak velocity index in the aorta and mean diastolic blood pressure rose in the lower pretransfusion haematocrit group.^24^ Even clinically stable preterm infants may exhibit normal left ventricular systolic function but show altered diastolic function.^25^ Ultimately, increasing HR after transfusion may indicate TACO in some preterm infants as the myocardium of the preterm infant has a low ability to respond to volume stress.^26, 27^ Such harm could be currently underestimated in this patient population. A recent analysis of systemic hemodynamic changes by electrical velocimetry detected changes in cardiac contractibility, blood flow, vascular resistance and fluid status temporally associated with RBC transfusion, but all parameters remained within the normal physiological range.^28^ These measurements used a minute-by-minute approach in a small ELGAN cohort, suggesting our high-granularity (0.017 to 1 Hz) and larger multicentre cohort may provide higher sensitivity.

Based on current understanding of the cause for anaemia in very preterm infants, we defined two periods of transfusion needs: (a) early anaemia, within the first 2 weeks after birth, most likely caused by blood loss (mostly iatrogenic);^29^ late anaemia from ≥3 weeks onwards, mostly characterized as hyporegenerative anaemia.^30^ Importantly, we found that the portion of preterm infants responding with increased HR is higher in early anaemia (postnatal age <2 weeks).

We also observed that approximately another 20% of infants responded with a decrease in HR after transfusion. Lower HR may indicate adequate time for ventricular filling and ejection.^24^ Interestingly, most infants exhibited no changes in cardiorespiratory parameters following transfusion. This suggests an adaptation of the myocardium to anaemia, especially beyond the second week of life, which is in line with the observation that in late anaemia echocardiographic parameters do not improve within 48 hours after transfusion.^24^ Combining our continuous vital signs approach with more closely monitoring of cardiac function is an important direction for future research. We observed similar percentages of preterm infants that exhibited increased or decreased RR and/or SpO2 following transfusion. However, data on simultaneously recorded levels of inspired oxygen were not available, making interpretation of these results complicated. Including other methods, such as near-infrared spectroscopy (NIRS), echocardiography or vascular videomicroscopy for perfusion indices, could serve as supplements to continuous cardiorespiratory monitoring in future patient blood management. Moreover, considering co-morbidities and medication with impact on the results will be an essential point for future work.

Using EHD offers considerable promise for assessing the clinical effects of therapeutic interventions, such as transfusions, in very preterm infants. Differences in data storage, access and sampling, as well as medico-legal regulatory processes in the respective centres and countries, can present challenges. Moreover, combining data across different centres presents regulatory (data sharing) and data harmonisation (differences in methodology) challenges. To address this, only de-identified, pre- processed data were combined, with each centre processing its own data locally to form a federated dataset. However, given the fact that the proportion of infants who exhibit a change in cardiorespiratory parameters is small, combining data across multiple centres is crucial. At one site data is not continuously stored and data collection had to be performed prospectively. This limited the number of infants included in the study but enabled greater control over data quality and additional parameters (apnoea rate) could be assessed. In contrast, whilst retrospective data collection can have limitations due to lower granularity of data (depending on the monitoring or data storage system),^31^ it can nevertheless substantially increase the number of participants.

## CONCLUSIONS

Our study highlights the promise of using highly-granular electronic monitoring data of cardiorespiratory parameters to describe the impact of transfusion practice in preterm infants. Our methodology may apply to investigate the effects of other therapeutic interventions and haemovigilance in very preterm infants. Our findings suggest the important need to better understand changes in cardiorespiratory parameters, as well as the clinical stability following RBC transfusion in preterm infants. The ultimate goal is to inform personalized patient blood management strategies beyond decision making by transfusion thresholds.

## CONTRIBUTORS

Conceptualization: SZ, SS, CD, ED; Data curation (retrospective): AH, IB, SZ, ED; Data collection (prospective): AS, MP; Data analysis: AH, TR, CZ, CH; Data interpretation: AH, TR, HvdS, SZ, SS, CH, CD, ED; Visualization: AH, TR, CH; Writing – first draft: CH, CD, ED; Writing – review and editing: All authors. All authors approved the final manuscript as submitted and agreed to be accountable for all aspects of the work.

## ETHICS APPROVAL

This study involves human participants. The study conformed to the Declaration of Helsinki and Good Clinical Practice and was approved, or granted a waiver of consent, by the responsible institutional review board or research ethics committee at each participating site (Berlin –EA2/096/25, no data used, waiver of consent; London - REC reference 25/HRA/0195, waiver of consent; Oxford - 23/NW/0155 and 19/LO/1085, written consent; Stockholm - Dnr 2022-06970-01, waiver of consent; see also per centre explanations in Supplemental Material).

## DATA AVAILABILITY STATEMENT

Imperial College London data: available subject to Imperial College Healthcare NHS Trust approval (ethical restrictions apply); requests to. Oxford data: due to ethical restrictions, the Oxford data set is available on request to. Karolinska University Hospital data: available subject to institutional and Swedish Ethical Review Authority approval (ethical restrictions apply); requests to. Analysis code is available at https://github.com/antoinehonore/neored-vs.

## OPEN ACCESS

For the purpose of open access, the authors have applied a CC BY public copyright licence to any Author Accepted Manuscript version arising from this submission.

## ABBREVIATIONS

BPD,: bronchopulmonary dysplasia;
CPAP,: continuous positive airway pressure;
EHR,: electronic health record;
HFO,: high-frequency oscillation;
HR,: heart rate;
IBI,: interbeat interval;
NEC,: necrotizing enterocolitis;
NICU,: neonatal intensive care unit;
NIRS,: near-infrared spectroscopy;
PMA,: postmenstrual age;
PNA,: postnatal age;
RBC,: red blood cell;
ROP,: retinopathy of prematurity;
RR,: respiratory rate;
SIMV,: synchronized intermittent mandatory ventilation;
SIPPV,: synchronized intermittent positive pressure ventilation;
SpO2,: peripheral oxygen saturation;
TACO,: transfusion- associated circulatory overload;
TRAGI,: transfusion-related acute gut injury;
TRALI,: transfusion- related acute lung injury

## ACKNOWLEDGEMENTS

Members of the Neonatal Transfusion Group include: Christof Dame^1,2^, Emöke Deschmann^3,4^, Suzanne F. Fustolo-Gunnink^5,6,7^, Lisanne Heeger^5,6^, Antoine Honoré^8^, Nina A M Houben^5,6^, Anne M Kelly^9,10^, Enrico Lopriore^5^, Helen V New^9^, Genny Raffaeli^11^, Till J. Rech^1^, Nora J. Reibel-Georgi^1^, Charles C. Roehr^12-14^, Simon J. Stanworth^9,15^, Alexandra Scrivens^16^, Hilde van der Staaij^5,6^. · Department of Neonatology, Charité – Universitätsmedizin Berlin, Berlin, Germany; 2 Department of Neonatology, Pediatric Pneumology and Neuropediatrics, Universitätsklinikum Schleswig-Holstein, Campus Kiel, Kiel, Germany; 3 Department of Women’s and Children’s Health, Karolinska Institutet, Stockholm, Sweden; 4 Department of Neonatal Medicine, Karolinska University Hospital, Stockholm, Sweden; 5 Division of Neonatology, Willem-Alexander Children’s Hospital, Leiden University Medical Center, Leiden, the Netherlands; 6 Sanquin Research, Sanquin Blood Supply Foundation, Amsterdam, the Netherlands; 7 Pediatric Hematology, Emma Children’s Hospital, Amsterdam University Medical Center, University of Amsterdam, Amsterdam, the Netherlands; 8 Department of Women’s and Children’s Health, Karolinska Institutet, Stockholm, Sweden; 9 Great Ormond Street Hospital, Haemophilia Comprehensive Care, London, United Kingdom; 10 NHS Blood and Transplant, United Kingdom; 11 Neonatal Intensive Care Unit, Fondazione IRCCS Ca’ Granda Ospedale Maggiore Policlinico, Milan, Italy; 12 National Perinatal Epidemiology Unit, Nuffield Department of Women’s and Reproductive Health, University of Oxford, Oxford, United Kingdom; 13 Faculty of Health and Life Sciences, University of Bristol, Bristol, United Kingdom; 14 Women’s and Children’s Division, Southmead Hospital, North Bristol NHS Trust, Bristol, United Kingdom; 15 Radcliffe Department of Medicine, University of Oxford, Oxford, United Kingdom; 16 Newborn Care Unit, Oxford University Hospitals NHS Foundation Trust, Oxford, UK.

### Acknowledgements

See Acknowledgements section for the full list of Neonatal Transfusion Network members.

## Data availability

Data is generally available upon request under ethical restrictions differing by centre; see Data Availability Statement for full details and contacts.

## Author Contribution(s)

See Contributors statement below for the full breakdown of author roles.

