## Supplemental Material for "Electronic health data exploring cardiorespiratory responses of transfusions in preterm infants: An international multicenter cohort study"

* Contributed equally as co-first authors

[#] Contributed equally as co-senior authors

[§] corresponding author

### Data Flows and Monitoring system information

#### Figure S1.1 Data flow chart at the Charité – Universitätsmedizin Berlin

At Charité – Universitätsmedizin Berlin data collection was retrospective. Approval has been given by the Institutional Review Board of the Charité (EA2/096/25). Data from the EPR system is fed continuously to the *health data platform (hdp)* and is then made available upon request as a data repository on a research server. The *hdp* data contains full laboratory, imaging, medication and clinical data. Vital signs are fed from the Monitoring system to the EPR as well but are resampled at varying frequencies of 1/min to 1/15min (0,01667 to 0,00111 Hz). Therefore, the vital sign data from the EPR could not be used for this analysis. To overcome this limitation, a dedicated data warehouse (*Philips Data Warehouse Connect*) was established recently and has been operational for the neonatal wards since early 2026. The warehouse is connected directly to the monitoring system consisting of Philips IntelliVue MX750 and MX800 Patient Monitors and stores raw monitoring data. The process to export data from this data warehouse to a research data repository for anonymized analysis has become operational in a beta version in mid 2026. Linking monitoring data to the correct patient record in the EHR depends on staff registering the patient in the monitoring system at admission; standard operating procedure on this registration workflow had to be finalised and has so far been adopted inconsistently across wards. The Institutional Review Board's waiver of consent for this retrospective design can only be granted on a retrospective submission.

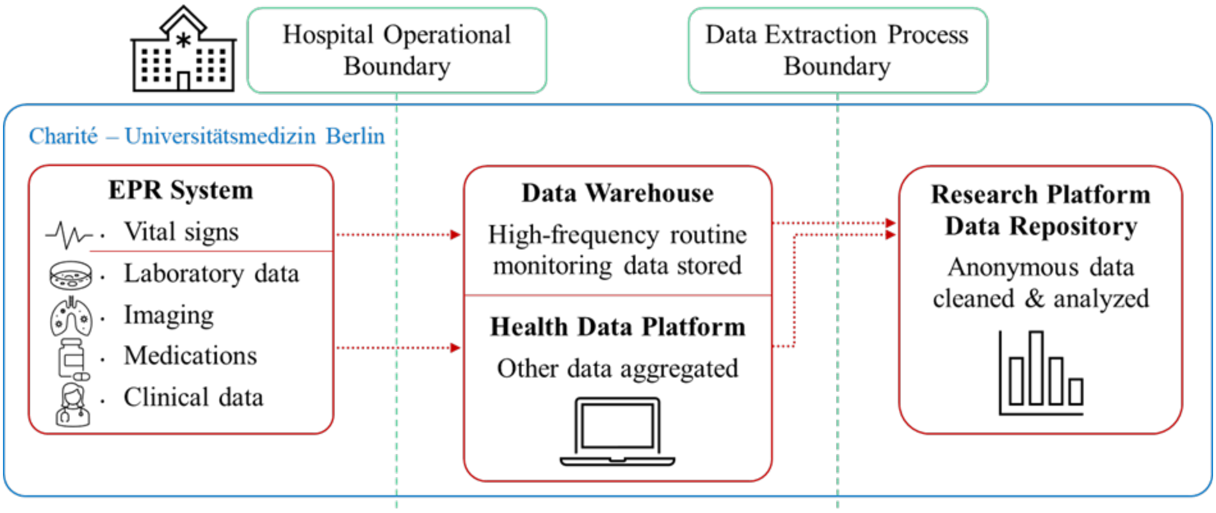

#### Figure S1.2 Data flow chart at the Imperial College London

At the Imperial College London study center, data was collected retrospectively from a centralized data warehouse for all infants admitted to Imperial College London NHS Trust tertiary NICU between 1st February 2023 and the first of February 2024. The HRA UK (Health Research Authority UK) has granted a waiver of consent for anonymized data (REC reference 25/HRA/0195). The vital sign recordings were downloaded from the electronic patient records (EPR) database, as well as the demographic information, laboratory data, the number and the type of transfusion and pre- and post- transfusion Hb. The duration and volume of transfusion as well as correct timings were extracted from a separate database used for electronic prescribing. High frequency vital signs data from Phillips Intellivue MX800 patient monitor system was sampled from a local Phillips Data Warehouse. The pre-processed monitor data re-sampled at 1Hz were used in the analysis.

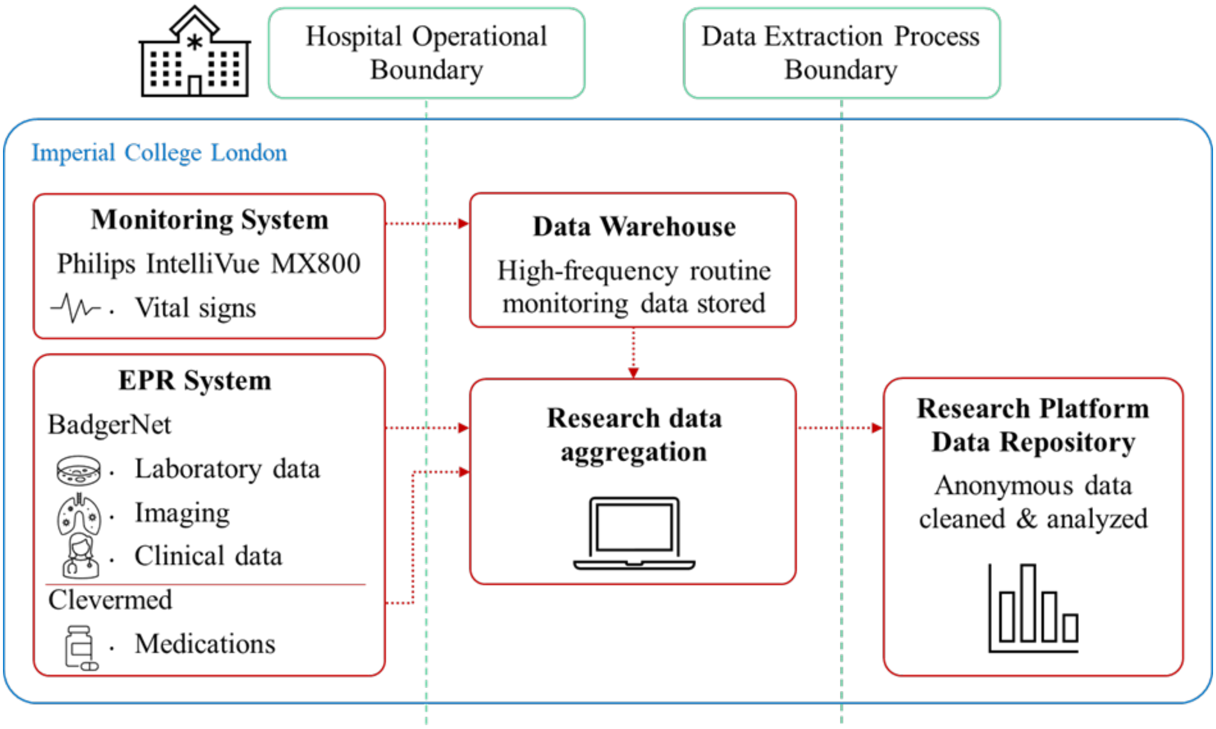

**Figure S1.3 Data flow chart at John Radcliffe Hospital and University of Oxford.**

At the Oxford study center, data was collected as a prospective cohort study conducted on the Newborn Care Unit, John Radcliffe Hospital, Oxford University Hospitals NHS Foundation Trust, Oxford, UK between 17th October 2023 and 5th October 2024. Additionally, 4 infants who had their vital signs recorded as part of another research study and also had a transfusion (between 5th December 2019 and 15th December 2024) were included in the analysis. The studies were approved by the UK National Research Ethics Service (references: 23/NW/0155, 19/LO/1085). Eligible families were given verbal and written information about the study, and written parental consent was obtained before inclusion in the study. A total of 24 infants were recruited to the study; 7 infants were excluded, leaving a total of 17 infants with 23 transfusion events included in the analysis.

Vital signs (heart rate, respiratory rate, oxygen saturation) were monitored using Phillips IntelliVue MX800 or MX750 monitors and data were continuously downloaded from the vital signs monitor using an electronic data capture software (iXtrend, ixitos, Germany) onto a research laptop for at least 12 hours before and after the start of the transfusion. Heart rate, oxygen saturation and respiratory rate (calculated by the monitor) were downloaded onto the laptop at a sampling rate of 0.97 Hz; the electrocardiograph (ECG, to measure heart rate) was measured with three electrodes placed on the infant's chest and recorded at 250 Hz and; the impedance pneumography (IP, to measure respiration) was measured using the same chest electrodes and recorded at a sampling rate of 62.5 Hz and, the photoplethysmography (PPG, to measure oxygen saturation and pulse) was measured using from a probe placed on the infant's foot or hand and recorded at 125 Hz. The start and end times of the transfusions were annotated post hoc onto the recordings using the times from electronic patient records (EPR), which are automatically uploaded when the clinical team scanned the blood packet to be transfused.

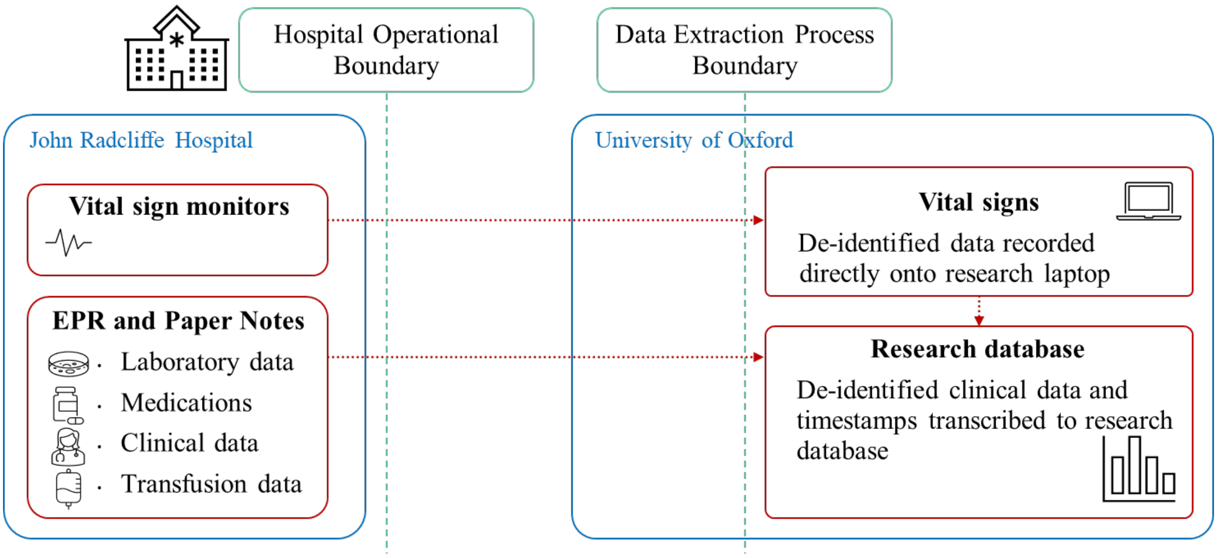

#### Figure S1.4 Data flow chart at the Karolinska Institutet Stockholm

The electronic health records (EHR) data of all infants hospitalized at Karolinska University Hospital between 2018 and 2023 was collected retrospectively from a centralized data warehouse. A waiver of consent was granted for the study by the Swedish Ethical Review Authority (Dnr 2022-06970-01).

From the EHR patient demographics information, all blood product type, transfusion dose and rate information, as well as hemoglobin level measurements were extracted. The vital signs recordings were collected from a separate database. We retrieved high frequency data from Philips IntelliVue MX800 Patient Monitor (Philips Healthcare, Amsterdam, Netherlands) from a local Philips Data Warehouse Connect database. We used the pre-processed monitor data resampled at 1Hz as basis signals for analysis.

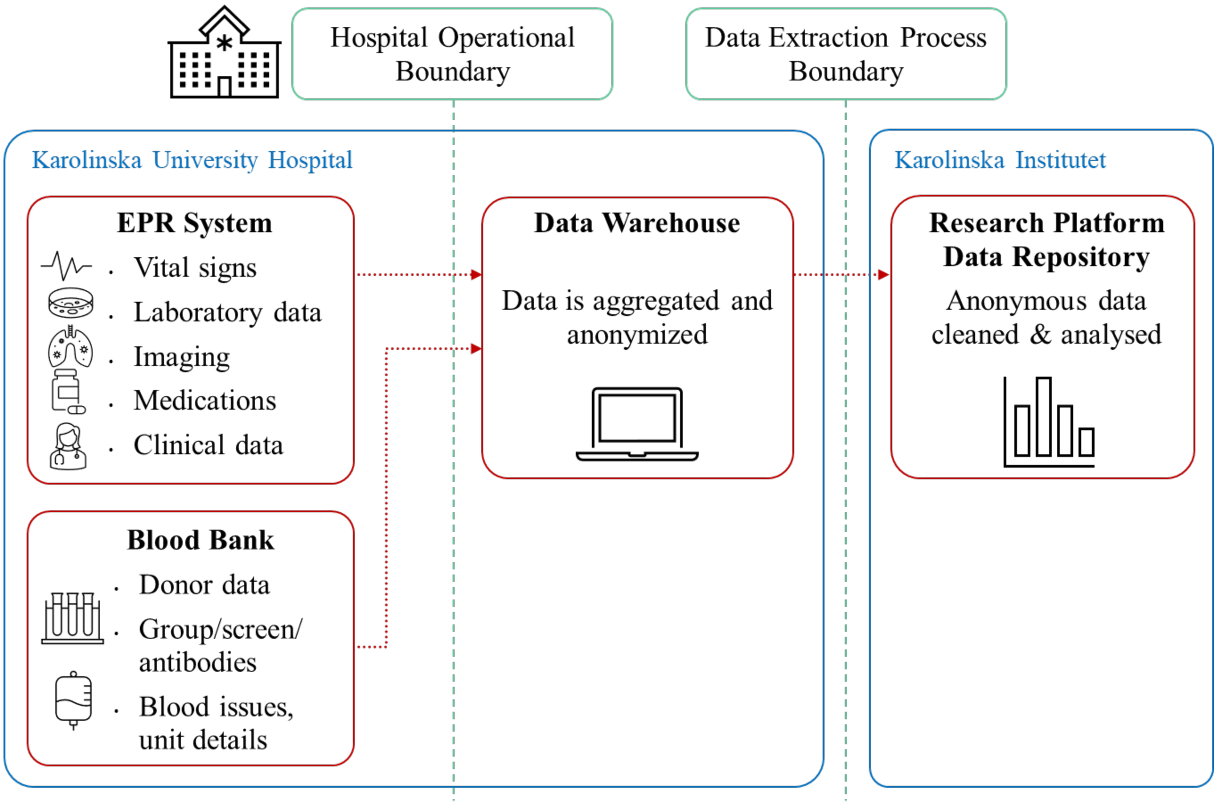

**Figure S2  Example vital signs from two different infants showing the analysis steps.**

Column 1 – Raw signals (blue) recorded directly from the infant's vital signs monitor. Column 2 – for each raw signal we investigated the average by calculating the mean in 1 hour time intervals and the variability by calculating the standard deviation in 1 hour time intervals. Column 3 – the change in each parameter from baseline was calculated by subtracting the mean of that parameter during the baseline period. In all columns, the grey shaded region indicates the transfusion period, and the grey dashed vertical line indicates the start of the transfusion. In column 3, the dashed black horizontal lines indicate the threshold values used to identify whether the change in vital signs was significant.

(A) Heart rate from one infant. The average heart rate in this infant increased significantly following the start of the transfusion. Heart rate variability initially increased and then decreased approximately 4 hours after the start of the transfusion, before returning to baseline. (B) Oxygen saturation from one infant. Average oxygen saturation decreased slightly during transfusion. Oxygen saturation variability decreases significantly from approximately 7 hours after the start of the transfusion.

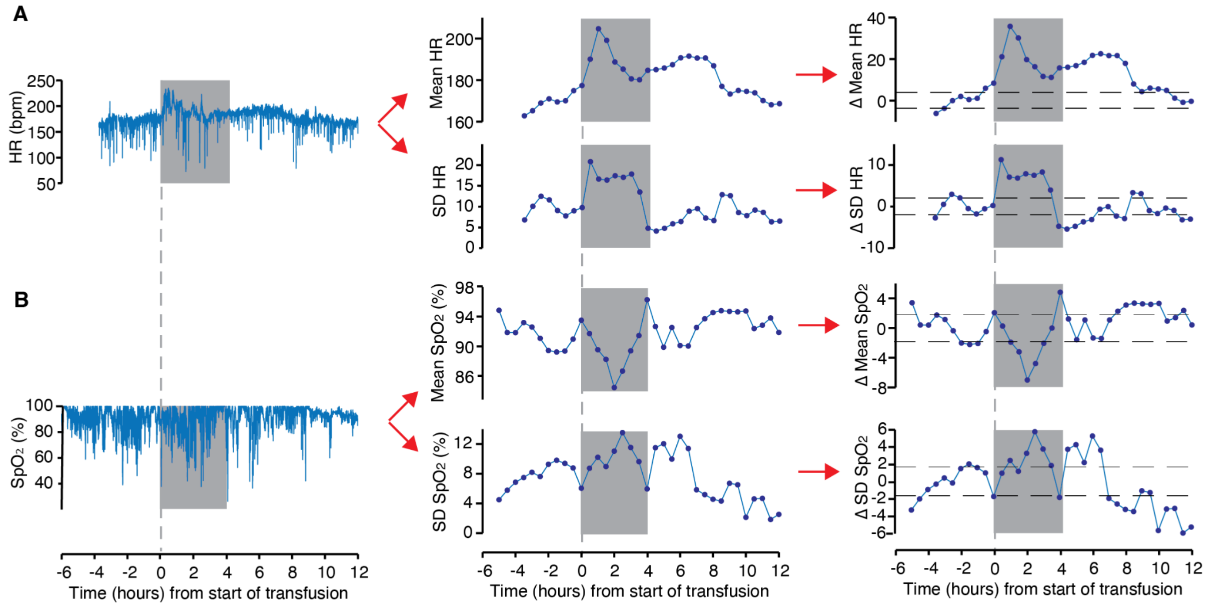

### Table S1  Monitoring system information at each centre

| | Berlin | London | Oxford | Stockholm |
|---|---|---|---|---|
| **Study period** | N/A | 01/02/2023 – 01/02/2024 | 11/2019 – 06/2024 | 2018–2023 |
| **Data collection** | Retrospective | Retrospective | Prospective | Retrospective |
| **Patient data management system (PDMS)** | COPRA / SAP | Clevermed/Badgernet | Cerner EPR and paper records | Clinisoft / TakeCare |
| **Monitoring system in the NICU** | Philips IntelliVue MX800 | Philips IntelliVue MX800 | Philips IntelliVue MX800 or MX750 | Philips IntelliVue MX800 |
| **Data granularity** | Pre-processed data at 1 Hz | Pre-processed data at 0.017 Hz | Raw signals up to 250 Hz and pre-processed at 1 Hz | Pre-processed data at 1 Hz |
| **Data storage system** | Philips Data Warehouse Connect | Philips Data Warehouse Connect | Research laptop connected to monitor (ixtrend software) | Philips Data Warehouse Connect |

### Transfusion-event definition and data cleaning

For each RBC-transfusion event, the start time was defined as the first of consecutive transfusion markers at most 1 hour apart, and the end time as the last volume part transfused. Volume (ml), patient prescription per current body weight (ml/kg body weight), rate (ml/h), and product specification were recorded, and the event dose was the sum of the doses of the individual transfusions.

In Stockholm data, when several rates were reported within an event, the median rate was used. After visual inspection of the resulting dose and rate distributions, outlier events (rate below 3 ml/h/kg or dose below 8 ml/kg) were removed.

In London, transfusion data were collected and plausibility-checked manually from the electronic health record, so no outlier removal was required.

### Respiratory-support modes

The respiratory-support modes encountered across centers were nasal high-flow, nasal continuous positive airway pressure (CPAP), BILEVEL/DuoPAP, synchronized intermittent positive-pressure ventilation (SIPPV)/synchronized intermittent mandatory ventilation (SIMV) with and without volume-targeted ventilation, and high-frequency oscillation (HFO; invasive or non-invasive). SIPPV/SIMV and invasive HFO were classed as mechanical ventilation.

### Significance thresholding

The threshold used to flag a significant post-transfusion change (one standard deviation of the baseline period, sustained for at least 6 consecutive time points, i.e. 3 hours or more) was chosen from visual assessment of the Oxford data set and was fixed before any analysis of the London or Stockholm data.

### Statistical analysis details

For cluster-based permutation testing, t-statistics at each post-transfusion time point were calculated by comparison with the pre-transfusion distribution; points exceeding the 97.5th percentile of the t-distribution were retained, temporally adjacent points were grouped into clusters, and cluster significance was assessed against 1000 permutations of the data using PALM software.

### Identification of cardiorespiratory events

Cardiorespiratory events were identified in data from Oxford and Stockholm only, where the sampling rate was sufficient for analysis. Apnoea rates were calculated at Oxford only where impedance pneumography (IP) was recorded, which provides accurate estimation of apnoeas compared with the respiratory rate signal derived on the monitors.[1]

Episodes of oxygen desaturation were identified from the peripheral oxygen saturation signal as periods during which oxygen saturation fell below 80% for at least 10 seconds. Episodes of bradycardia were identified as periods during which the heart rate fell below 100 beats per minute (bpm) for at least 15 seconds.

To calculate apnoea rates, IP signals were processed using an algorithm by Adjei et al. validated for the identification of inter-breath intervals (IBIs) and apnoea in infants.[1] Briefly, the algorithm first filters the signal to reduce the noise introduced by movements and cardiac activity. An adaptive amplitude threshold (set at 0.4 times the standard deviation of the signal over the preceding 15 breaths) is used to identify individual breaths. Finally, a support vector machine algorithm was applied to all potential episodes of apnoea (defined as IBIs longer than 15 seconds) to remove periods of low amplitude erroneously identified as apnoea due to noise or shallow breathing. This model was trained and validated on IP signals completely independent from our current data sample. Long IBIs marked as noise/shallow breathing were discarded from further analysis. Apnoea rate was calculated as the number of apnoeas (IBIs greater than 15 seconds) per hour.

### Results

#### Figures S3 Changes in cardiorespiratory metrics in the data collected from the different study centers.

Mean and standard deviation (SD) of the change in cardiorespiratory metrics before and after the start of RBC transfusion. Results are grouped for each study center according to transfusion events with no change in the corresponding cardiorespiratory metric (red), transfusion events with a decrease in the cardiorespiratory metric (blue), transfusion events with an increase in the cardiorespiratory metric (orange). Means are indicated by solid lines and the shaded area indicates the standard deviation. Black dashed vertical line indicates the start of transfusion (time = 0).

#### Figure S3.1 Results at the Imperial College London

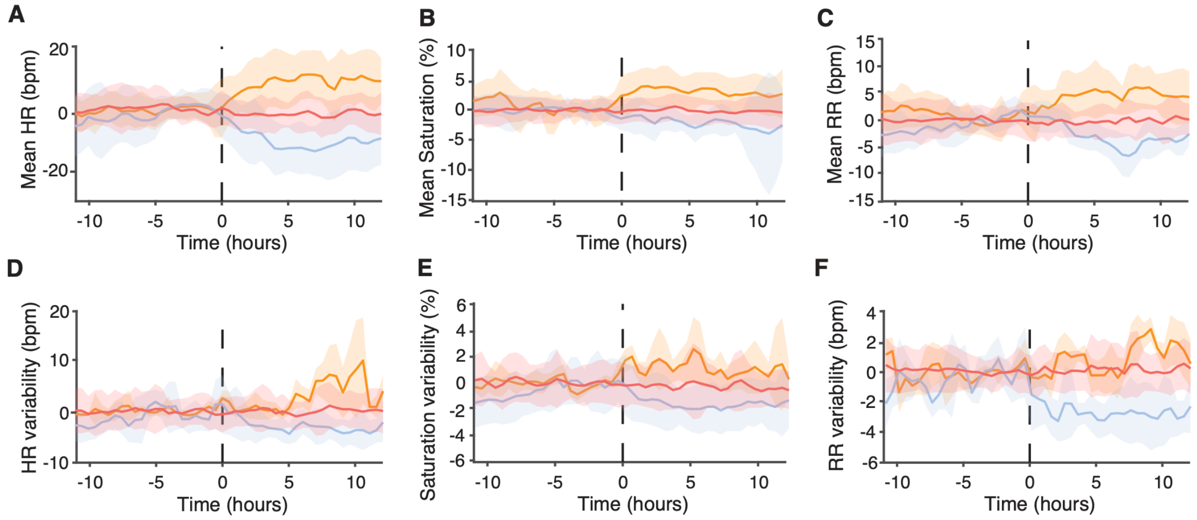


#### Figure S3.2 Results at the University of Oxford

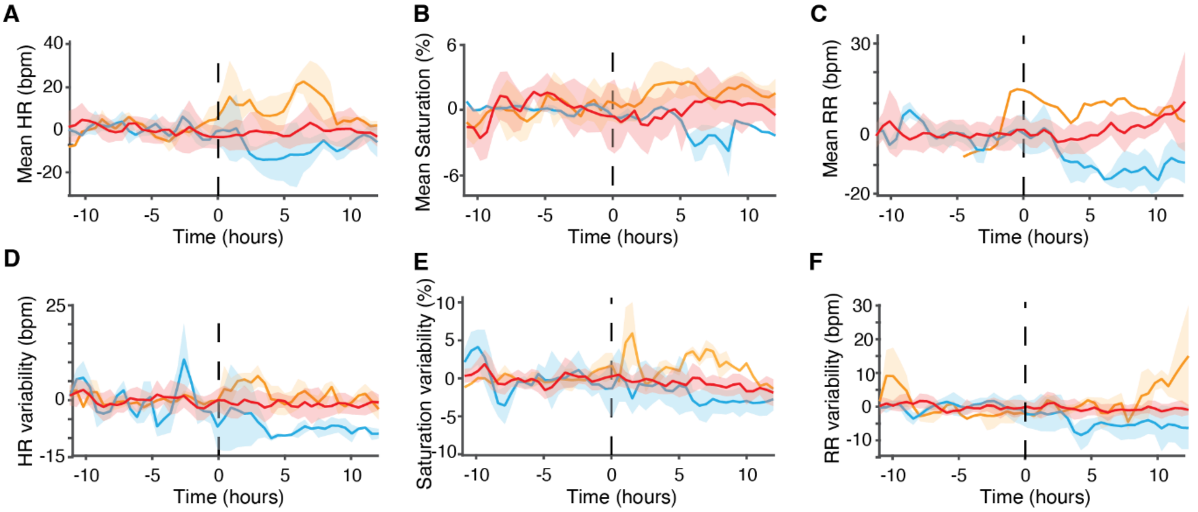

### Figure S3.3 Results at the Karolinska Institutet Stockholm

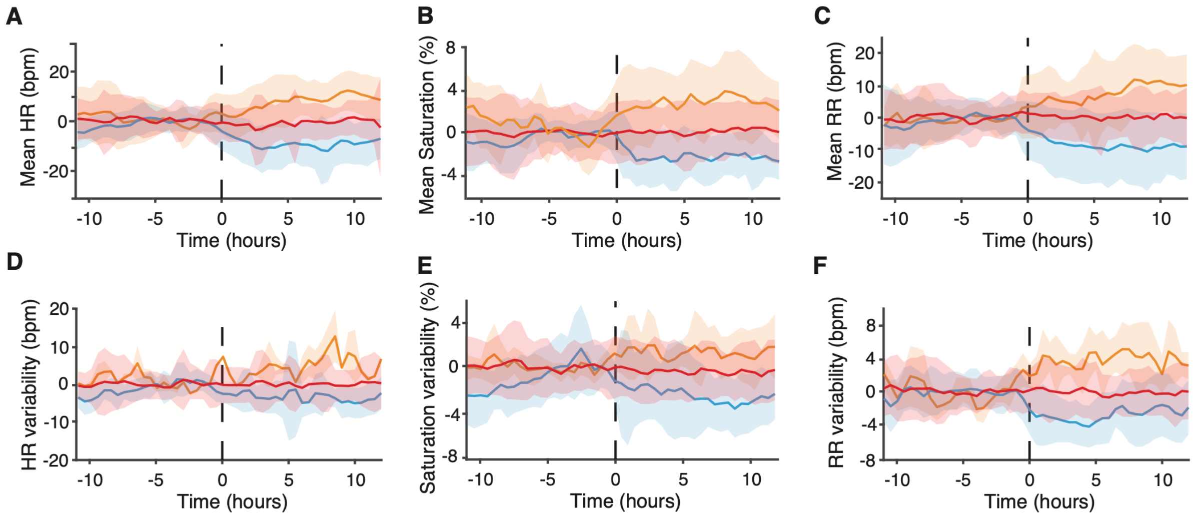
